# Plasma Proteomics Identifies a Microtesla Magnetic Therapy Response Signature in Long COVID

**DOI:** 10.64898/2026.08.25.26361319

**Authors:** Nathan R. Brady, Alexandra Canori, David Maltz, Douglas Y. Kirsher, Wenyu Zhou, Jacqueline Becker, David F. Putrino, Blake T. Gurfein

## Abstract

Cognitive impairment is a disabling feature of Long COVID with no established disease-modifying therapy, and little is known about the biological changes accompanying clinical improvement. Microtesla Magnetic Therapy (MMT) is a low amplitude radiofrequency electromagnetic field intervention delivered to the whole brain. In a randomized, sham-controlled feasibility trial, at home MMT was feasible, safe, and well tolerated, with evidence of clinical improvement among treated participants. We explored molecular changes associated with response using SomaScan 11K plasma proteomics on paired baseline and week 4 samples. Participants were classified post hoc within each treatment arm using a clinician-selected response phenotype integrating cognitive and symptom domains. These groups were used for proteomic, pathway, and OrganAge analyses. MMT response was associated with selective proteome remodeling and an exploratory 17 protein response pattern in which Hedgehog interacting protein (HHIP), a Hedgehog signaling antagonist, was most strongly associated with response. Directional pathway analysis identified patterns consistent with lower inflammatory and injury biology and higher repair and adaptive remodeling. OrganAge analysis showed trends toward lower Brain and Organismal OrganAge with MMT. These findings prioritize HHIP and the exploratory 17 protein response pattern for prospective validation and support evaluation of plasma proteomics for monitoring treatment response.

## INTRODUCTION

Long COVID, or post-acute sequelae of SARS-CoV-2 infection, is a chronic multisystem condition that can persist for months to years after acute infection. Cognitive impairment is among its most common and debilitating manifestations ^1,2^. Patients commonly report difficulties across cognitive domains, often accompanied by fatigue, post-exertional malaise, sleep disturbance, and mood symptoms. Together, these symptoms can substantially impair daily function, quality of life, and the ability to return to work ^3–6^.

Evidence suggests that cognitive impairment associated with Long COVID may reflect interacting systemic and neurobiological processes. Proposed mechanisms include viral persistence, persistent immune activation, vascular injury, complement and coagulation activation, blood-brain barrier disruption, mitochondrial stress, tissue injury, and neuroinflammatory signaling ^7–9^. Longitudinal plasma proteomics provides a noninvasive approach to characterize systemic molecular changes over time and examine their relationship to later clinical outcomes ^10,11^.

Microtesla Magnetic Therapy (MMT) is a low amplitude, nonthermal radiofrequency electromagnetic field intervention delivered through a head-worn device designed to provide whole brain exposure. Preclinical studies of transcranial microtesla magnetic fields show reductions in inflammation and oxidative stress together with neuroprotective effects, providing a rationale to examine whether MMT influences injury and repair biology ^12^. In the triple-blind, first-in-human, randomized, sham-controlled trial, at-home MMT was feasible, safe, and well tolerated in individuals with objective cognitive impairment related to Long COVID. Exploratory analyses showed nominally significant between-group differences favoring active treatment on selected cognitive measures and emotional well-being^13^.

The molecular changes accompanying MMT and their relationship to clinical response remain undefined. Cognitive impairment associated with Long COVID often involves executive function, processing speed, and sustained attention ^6,14^, but occurs within a broader syndrome that can include fatigue, post-exertional malaise, and mood disturbance. Treatment-related improvement may therefore differ across cognitive and symptom domains. A single endpoint may miss domain specific improvement, whereas an overly broad composite may obscure a focused response. Linking early molecular changes to an exploratory, clinically informed response framework may help identify biological patterns associated with multidomain improvement.

To investigate molecular changes associated with MMT, we conducted an embedded exploratory plasma proteomic analysis of the parent feasibility trial using the SomaScan 11K platform, an aptamer-based assay that measures more than 10,000 plasma protein targets ^15^. To our knowledge, this is the first randomized, sham-controlled human trial to use large scale plasma proteomics to characterize circulating protein changes during a therapeutic electromagnetic field intervention. Using paired baseline and week 4 profiles, we tested whether MMT was associated with selective changes in the plasma proteome, whether those changes differed by week 8 response status, and whether response associated proteins converged on coherent biological programs.

Using responder groups defined by the post hoc clinician-selected response phenotype for molecular comparisons, we identified HHIP as the leading protein associated with response among MMT participants and found pathway patterns consistent with lower inflammation and injury biology and higher repair and adaptive remodeling biology. We also applied OrganAge modeling as an exploratory systems level analysis ^16^.

## RESULTS

### Study design and plasma proteomic profiling workflow

This exploratory plasma proteomic analysis was embedded within a randomized, sham-controlled feasibility trial of MMT in participants with cognitive impairment associated with Long COVID. Paired week 0 and week 4 plasma samples from 30 participants, including 20 assigned to MMT and 10 assigned to sham, were profiled using SomaScan, with clinical outcomes assessed through week 8. Early plasma protein changes were compared across responder groups defined post hoc by the clinician-selected response phenotype. The workflow included paired SomaScan change analysis, characterization of the MMT-associated protein set and the exploratory 17 protein response pattern, HHIP prioritization, pathway analysis, and exploratory OrganAge modeling (Figure 1).

**Figure 1.**
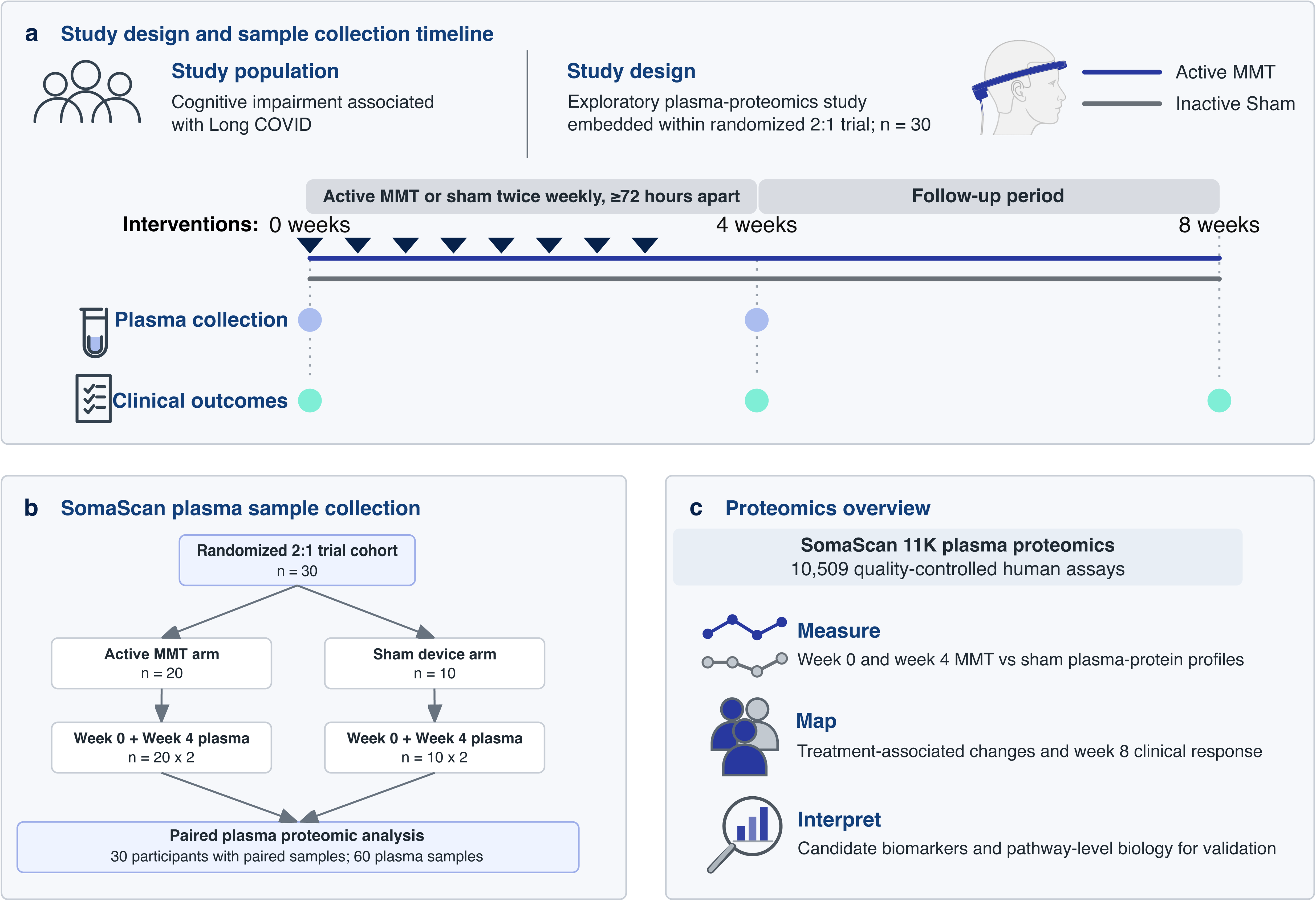
Study design and SomaScan workflow

### The post hoc clinician-selected response phenotype defines responder groups for exploratory molecular analysis

Clinical response was summarized using a post hoc, clinician-selected response phenotype developed specifically for this study. Following discussion of the available measures, the clinician recommended five components that covered a cross section of cognitive and symptom outcomes: processing speed, sustained attention, executive flexibility, working memory, and post exertional malaise. Table 1 summarizes the selected measures, their clinical domains, and the direction of improvement used in the molecular analyses.

Using the week 8 2-of-5 response rule, 16 of 20 MMT-treated participants met criteria for response, compared with 3 of 10 sham-treated participants (two-sided Fisher’s exact p = 0.0147; Figure 2a), corresponding to an observed risk difference of 50 percentage points.

**Figure 2.**
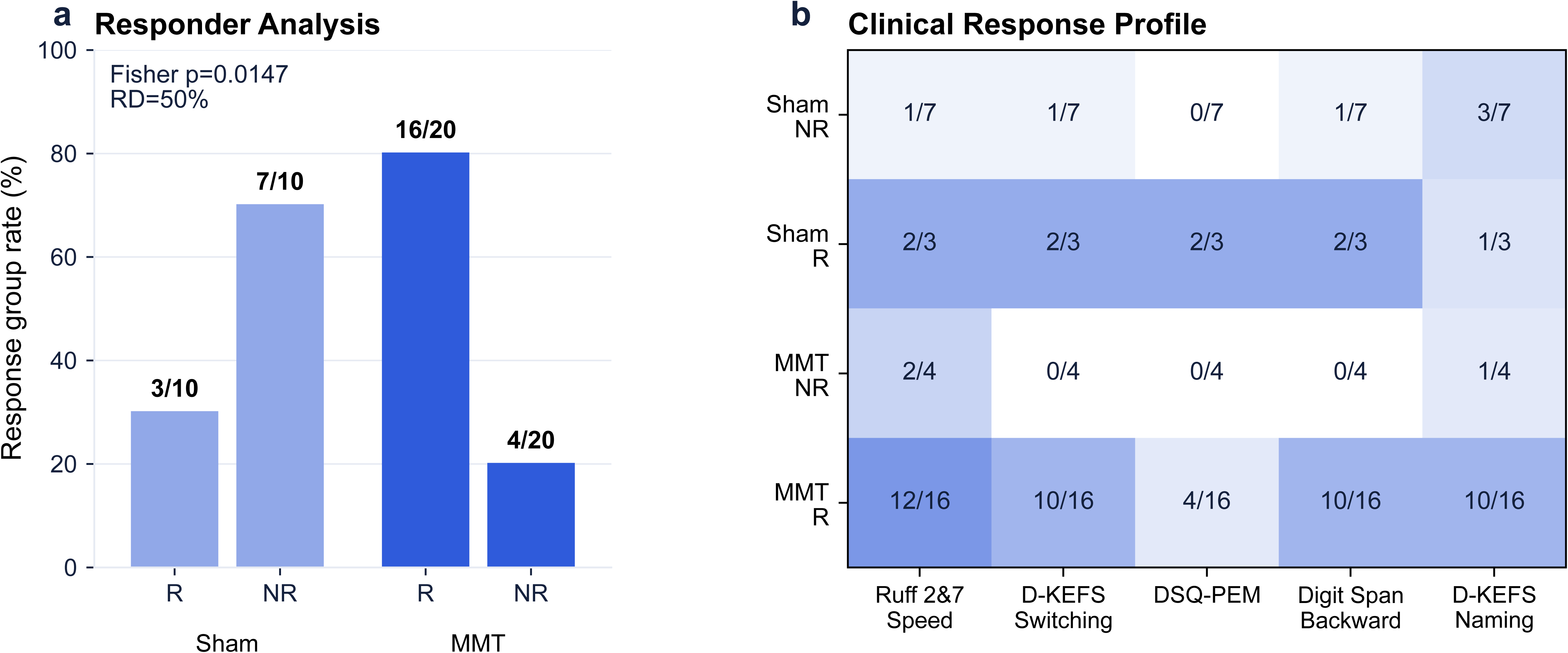
Post hoc clinician-selected response phenotype

Among MMT responders, component improvement criteria were met most often for Ruff 2&7 Controlled Speed, D-KEFS Category Switching, WAIS-IV Digit Span Backward, and D-KEFS Color Naming. Improvement on the DePaul Symptom Questionnaire–Post-Exertional Malaise (DSQ-PEM) occurred in a smaller subset (Figure 2b; Supplementary Figure 1).

### Selective plasma proteome changes associated with MMT

Principal component analysis of the SomaScan profiles showed substantial overlap among baseline Long COVID, sham week 4, and MMT week 4 samples in the first two principal components (Figure 3a). MMT week 4 samples showed somewhat broader dispersion, but the PCA did not show clear global separation by treatment group or time point.

**Figure 3.**
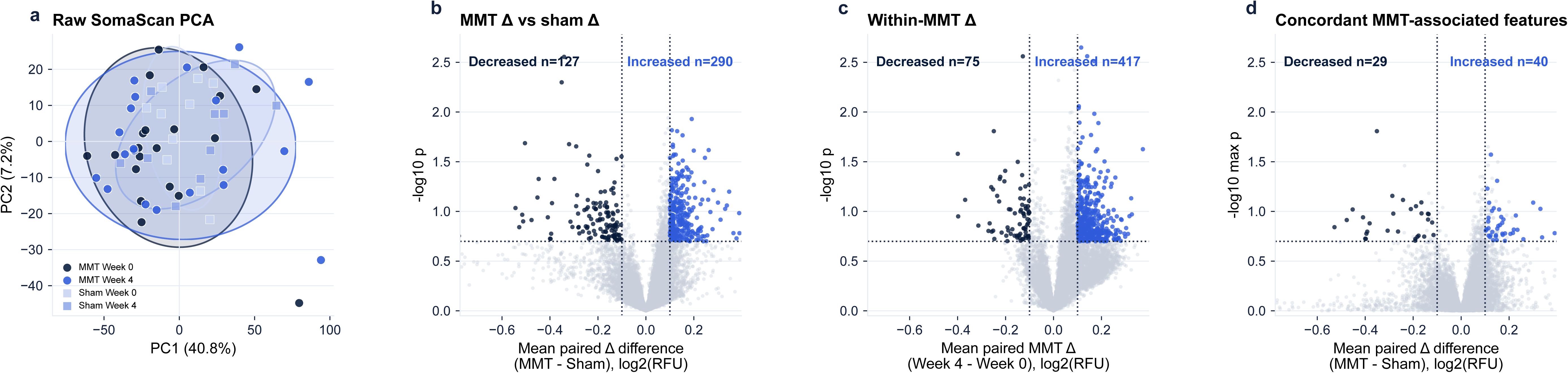
Exploratory SomaScan feature-screening strategy

Paired week 0 to week 4 SomaScan changes were analyzed using two exploratory screens. One screen compared longitudinal changes between MMT and sham; the other identified changes from baseline within the MMT group. Across the two screens, 69 SomaScan features corresponding to 66 unique protein targets met both criteria and showed matching directions of change (Figure 3b–d). The set contained both MMT-associated increases and decreases, consistent with selective rather than global plasma proteome remodeling.

### Exploratory 17 protein response pattern

The concordant MMT-associated set was intersected with the MMT responder versus nonresponder analysis, yielding the exploratory 17 protein response pattern.

To summarize the exploratory 17 protein response pattern at the participant level, we calculated a direction aligned composite score. Protein changes were standardized and oriented so that values in the MMT responder direction contributed positively. Higher scores therefore indicate greater similarity to the molecular pattern observed in MMT responders.

The composite score for the exploratory 17 protein response pattern was higher in MMT responders than in MMT nonresponders (p = 0.00289), sham responders (p = 0.00413), and sham nonresponders (p = 0.00460; Figure 4b). Thus, the exploratory 17 protein response pattern was most strongly expressed in MMT responders.

**Figure 4.**
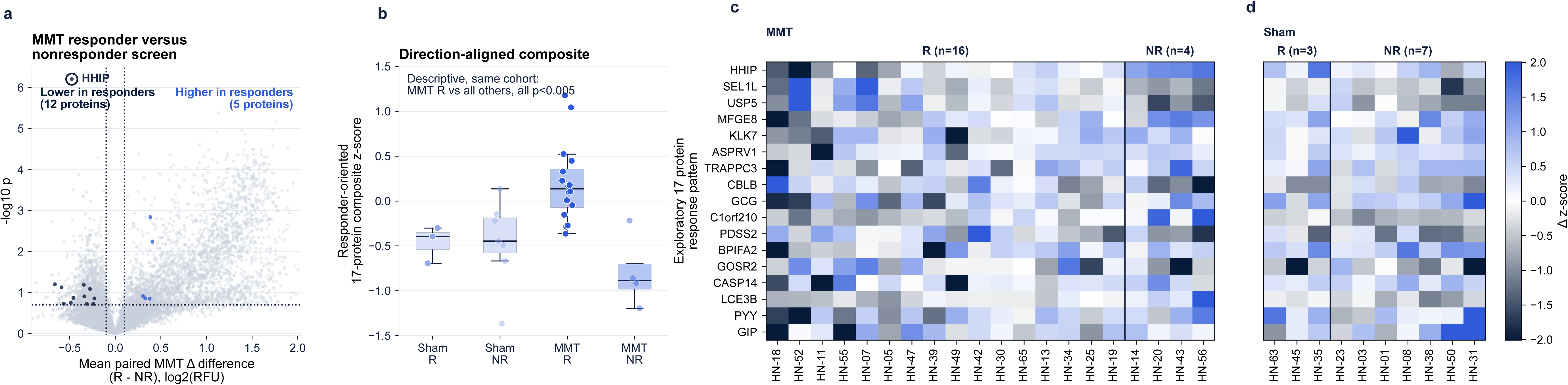
Exploratory 17 protein response pattern

Heatmaps were generated from standardized week 0 to week 4 protein changes without direction alignment, preserving the observed direction of change for each protein in the MMT and sham groups (Figure 4c,d). The heatmaps showed a visually more consistent pattern among MMT responders than among MMT nonresponders and sham participants. The composite score summarizes similarity to the MMT responder pattern, whereas the heatmaps display the individual protein changes contributing to that summary.

Supplementary Figure 2 shows raw week 0 and week 4 SomaScan trajectories for the 17 proteins in MMT participants. Thin lines show paired participant values, and thicker lines show group mean trajectories for MMT responders and nonresponders. The plots preserve each protein’s observed direction of change before direction alignment for the composite. Several proteins showed different mean trajectories between responder groups, including HHIP, which decreased in MMT responders and increased in nonresponders.

Together, the composite score, heatmaps, and raw trajectories show that the exploratory 17 protein response pattern was most evident in MMT responders.

### HHIP is the leading protein associated with response among MMT participants

Among the individual responder associated proteins, HHIP showed the strongest separation between MMT responders and nonresponders. HHIP change was defined as week 4 minus week 0 and was lower in MMT responders than in nonresponders (responder minus nonresponder difference = −0.477 log2 RFU; full screen q = 0.0066; exact two-sided permutation p = 0.000413; Figure 5a). The corresponding sham responder comparison was not significant (p = 0.29).

**Figure 5.**
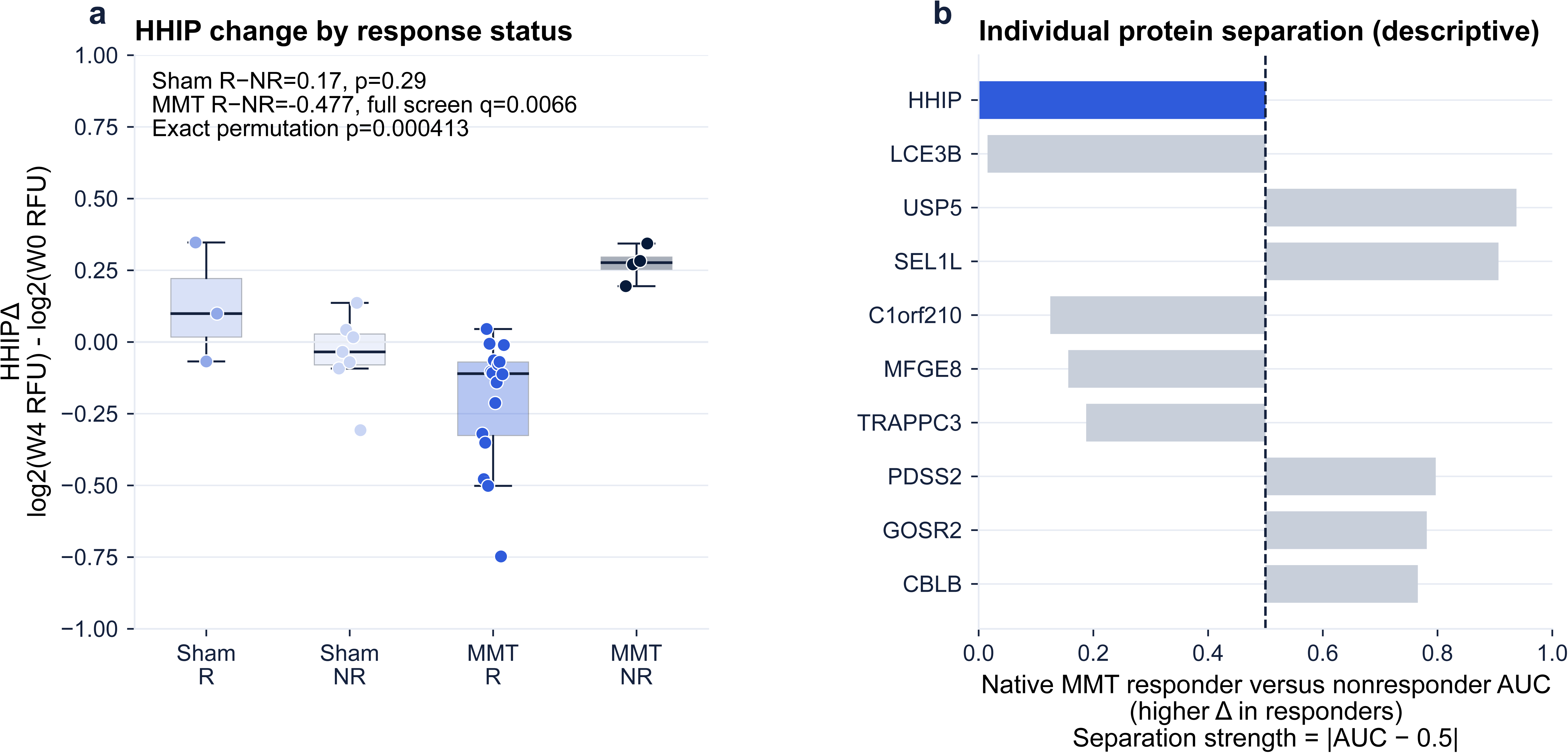
HHIP is the leading protein associated with response among MMT participants

Among the 17 proteins, HHIP ranked first by responder versus nonresponder separation (Figure 5b). Lower HHIP change completely separated the 16 MMT responders from the 4 nonresponders (native AUC = 0.0; AUC distance from 0.5 = 0.5), and this ordering was retained in bootstrap and leave one participant out analyses. Because HHIP was selected and evaluated in the same small cohort, these analyses support its prioritization for further study but do not establish monitoring or predictive performance. A targeted HHIP ELISA provided supporting evidence: HHIP decreased in 10 of 14 displayed MMT responders and 2 of 4 nonresponders (Supplementary Figure 4).

To explore a possible interpretation of the HHIP dynamics associated with MMT, we developed an unfitted conceptual model of HHIP/SHH inflammation resolution (Supplementary Figure 3). The model asks how repeated MMT-associated reductions in circulating HHIP might create transient SHH permissive windows; it does not reproduce participant biology. It contains three normalized states—circulating HHIP, unresolved damage, and inflammation—with effective SHH/Hedgehog signaling and repair (1 − damage) as algebraic readouts. One conceptual week with three MMT sessions transiently lowers HHIP and increases SHH permissiveness. Under the model assumptions, the four week simulation showed greater reductions in unresolved damage and inflammation than the one week simulation. These trajectories are a framework for understanding possible MMT HHIP dynamics, not estimates of efficacy, participant responses, or pathway engagement.

### Directional overrepresentation analysis highlights inflammation, injury, and repair related patterns

The directional overrepresentation analysis (ORA) examined whether protein sets associated with MMT and clinical response converged on broader biological programs. Results were consistent with two patterns: lower inflammation and injury biology and higher repair and adaptive remodeling biology (Figure 6a). Ten inflammation and injury programs met the FDR threshold, with the strongest ORA evidence for innate and myeloid inflammation (best FDR q = 0.000461). Ten repair and adaptive remodeling programs met the FDR threshold, with the strongest evidence for membrane trafficking and vesicle transport (best FDR q = 1.87 × 10^−8^).

**Figure 6.**
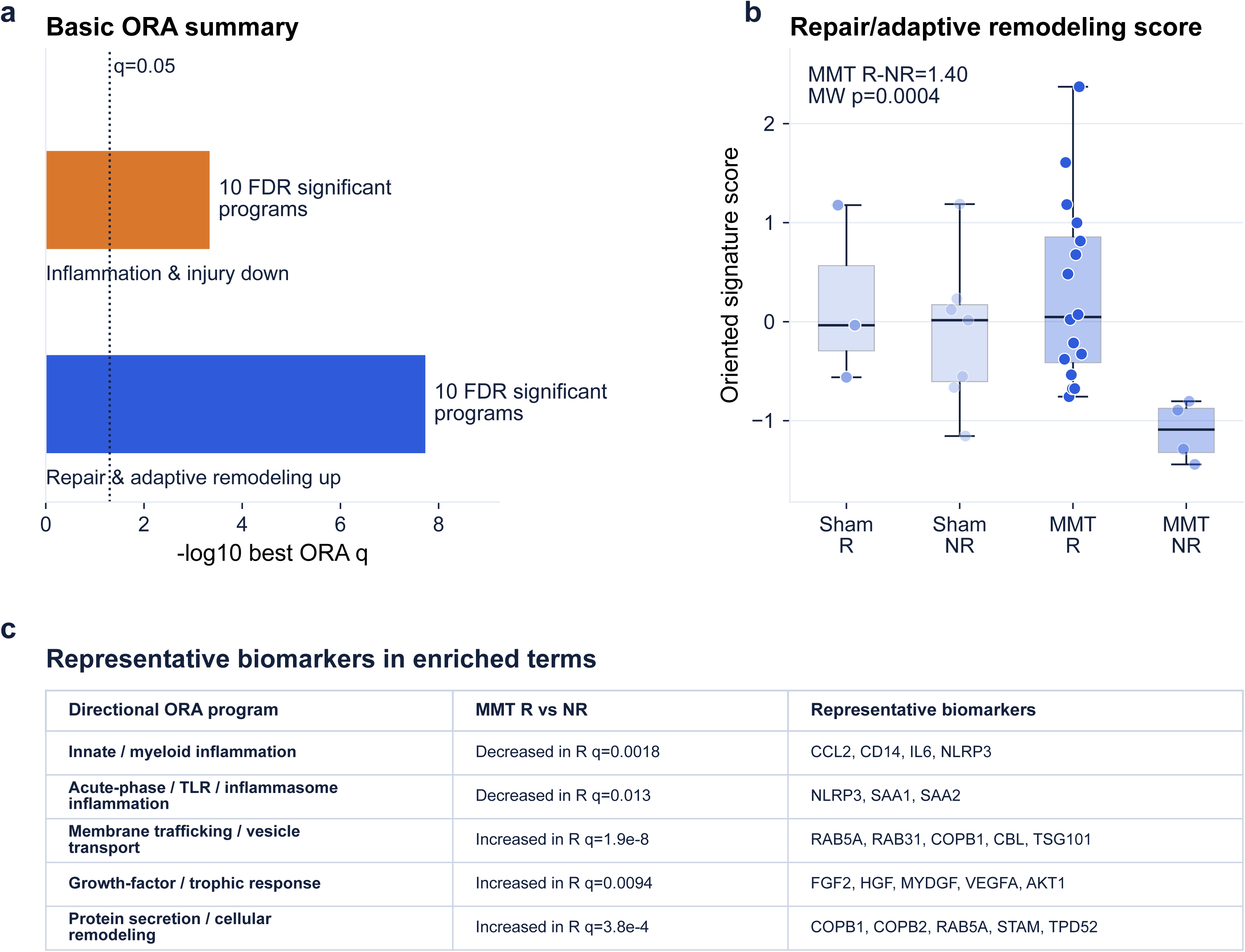
Directional ORA summary associated with response

We next asked whether participant level protein program scores were associated with response. Higher scores indicated greater alignment with the lower inflammation and injury pattern or the higher repair and adaptive remodeling pattern. Within MMT, responders had higher scores for the lower inflammation and injury pattern than nonresponders (Mann Whitney p = 0.00743; q = 0.0223) and for repair and adaptive remodeling (p = 0.000413; q = 0.00186; Figure 6b). Programs contributing to these patterns included lower innate and myeloid inflammation and lower acute phase, TLR, and inflammasome inflammation, together with higher membrane trafficking and vesicle transport, growth factor and trophic response, and protein secretion and cellular remodeling (Figure 6c). Detailed ORA results and ranked enrichment sensitivity are shown in Supplementary Figures 5 and 6.

Together, Figure 6 is consistent with an association between response status and a plasma protein pattern of lower inflammation and injury biology and higher repair and adaptive remodeling biology. Supplementary Figure 5 shows term level ORA results, and Supplementary Figure 6 shows ranked enrichment sensitivity.

### OrganAge analysis shows exploratory directional trends toward lower Brain and Organismal OrganAge with MMT

Given the multisystem nature of Long COVID and the relationship between inflammation and cellular aging, we applied pretrained plasma OrganAge models ^16^. These SomaScan models use organ enriched plasma protein signatures to estimate biological age gaps across 11 major organs. We evaluated changes in these signatures from week 0 to week 4. AgeGap values were analyzed as standardized paired changes, with negative values indicating a lower predicted biological age gap at week 4 than at baseline.

The OrganAge treatment screen showed negative MMT versus sham effects for several signatures, indicating lower week 4 minus week 0 OrganAge change in MMT than sham (Figure 7a). The strongest directionally lower effects were observed for Intestine (coefficient = −0.48; p = 0.062; q = 0.253), Liver (coefficient = −0.40; p = 0.071; q = 0.253), Brain (coefficient = −0.42; p = 0.094; q = 0.253; Figure 7b), Organismal (coefficient = −0.48; p = 0.105; q = 0.253; Figure 7c), and Muscle (coefficient = −0.48; p = 0.106; q = 0.253). Within MMT, responders showed directionally lower Brain OrganAge change than nonresponders (responder minus nonresponder = −0.24; Mann Whitney p = 0.750; Figure 7d). The Organismal difference was small (responder minus nonresponder = −0.02; p = 0.437; Figure 7e). Neither responder comparison was statistically significant.

**Figure 7.**
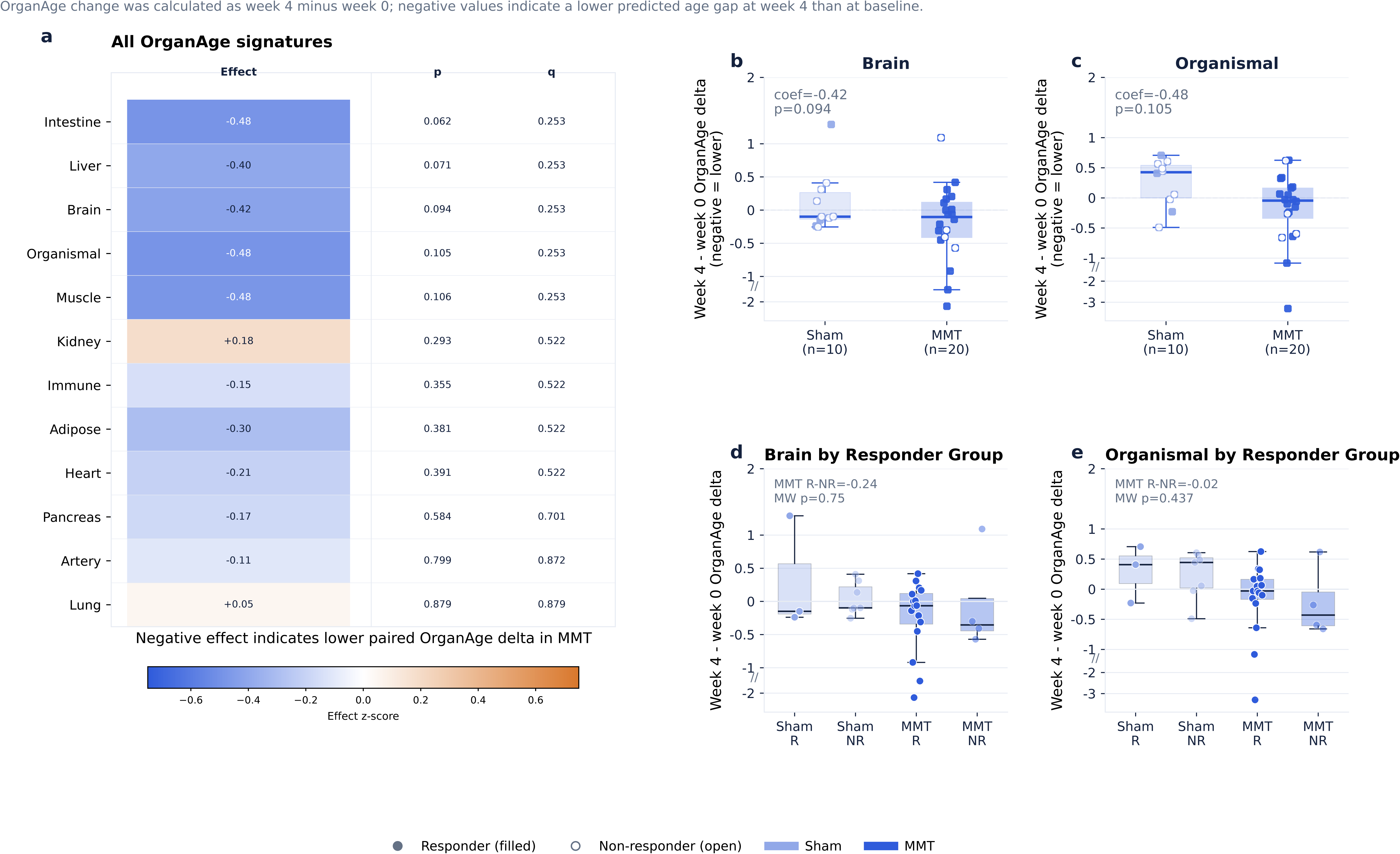
OrganAge responder analysis

These exploratory OrganAge findings provide directional systems-level context but do not establish treatment benefit. Lower estimated organ ages have been associated with lower disease and mortality risks and greater longevity in independent cohorts ^16,17^. Although lower OrganAge estimates do not establish organ rejuvenation or functional improvement, these findings provide a hypothesis-generating extension of the broader proteomic finding that beneficial clinical responses to MMT were linked to early molecular changes, including protein signatures associated with aging biology.

Overall, the above analyses linked MMT-associated clinical response to early, selective plasma-proteome remodeling rather than a broad proteome-wide shift. The post hoc response phenotype defined responder groups for molecular comparisons; SomaScan identified an exploratory 17 protein response pattern led by HHIP and associated with lower inflammation and injury biology and higher repair and adaptive remodeling biology. OrganAge analyses showed directionally lower Brain and Organismal changes in MMT than sham.

## DISCUSSION

This study links clinical response to early plasma-protein changes during MMT treatment for cognitive impairment associated with Long COVID. Clinical response was assessed using a clinician-selected response phenotype defined post hoc from our clinical study ^13^. Under the exploratory 2-of-5 definition, clinical response was more frequent with MMT than with sham. In this study, this phenotype was used to define responder groups for exploratory molecular comparisons.

The main molecular finding was selective plasma proteome remodeling associated with MMT rather than a broad proteome wide shift. The MMT responder pattern included lower inflammation and injury biology, higher repair and adaptive remodeling biology, and lower HHIP, a Hedgehog signaling inhibitor that sequesters Hedgehog ligands ^18–20^.

PCA showed overlapping baseline Long COVID, sham week 4, and MMT week 4 plasma profiles and did not indicate a broad global plasma proteome shift. SomaScan identified a selective MMT-associated protein set that included 17 proteins associated with response.

HHIP was the leading protein associated with response among MMT participants. HHIP decreased in MMT responders and separated MMT responders from nonresponders. Because HHIP is a Hedgehog ligand-sequestering feedback antagonist, the decrease observed among MMT responders is consistent with, but does not establish, a state more permissive for endogenous SHH/Hedgehog signaling ^21,22^. Across injury models, transient SHH/Hedgehog signaling has been linked to BBB stabilization and immune quiescence, GLI-dependent IL-10 induction, pro-repair macrophage programs, efferocytosis, wound healing, and vascular repair biology ^23–26^. ELISA measurement of HHIP provided supporting evidence for the SomaScan result, with HHIP decreasing more frequently in responders than in nonresponders. However, because HHIP was selected and re-evaluated in the same small discovery cohort, and because the ELISA subset was limited, these data do not establish HHIP as a validated standalone response biomarker or prove Hedgehog pathway activation in participants. Further testing in an independent cohort will be required to determine whether HHIP change has reproducible monitoring or predictive value.

The conceptual HHIP/SHH model in Supplementary Figure 3 was developed to help interpret the HHIP dynamics associated with MMT. The model assumes that transient increases in Hedgehog signaling reduce unresolved damage and inflammation, with remaining damage influencing later inflammation. Under the model assumptions, the four-week simulation showed greater reductions in unresolved damage and inflammation than the one-week simulation. Because the model is unfitted and prolonged Hedgehog activation can contribute to fibrotic or maladaptive remodeling, it should not be interpreted as evidence that reduced circulating HHIP is beneficial, that CNS SHH/SMO/GLI signaling was engaged, or that the simulated schedules estimate MMT efficacy ^27,28^.

Pathway level analysis provided broader biological context for the response associated protein pattern. ORA showed that response among MMT participants was associated with lower inflammation and injury biology and higher repair and adaptive remodeling biology. Within MMT, responders had higher program scores than nonresponders for both the lower inflammation and injury pattern and the higher repair and adaptive remodeling pattern. These patterns included programs involving innate and myeloid inflammation, acute phase, TLR, and inflammasome biology, membrane trafficking and vesicle transport, trophic response, secretion, and cellular remodeling. These programs overlap with biological processes reported in Long COVID, including immune activation, vascular injury, complement and coagulation, blood brain barrier disruption, mitochondrial stress, and tissue injury ^7–9^, and with preclinical MMT findings of lower inflammatory and oxidative stress signals ^12^. Together, the ORA results are consistent with lower inflammation and injury and higher repair biology among MMT responders.

OrganAge analysis provided a systems level extension of the plasma proteomic findings. Several OrganAge signatures showed directionally lower week 4 minus week 0 change in MMT than sham, including Brain and Organismal signatures, although the treatment comparisons did not meet FDR adjusted significance (Figure 7a). Brain and Organismal OrganAge changes were also directionally lower in MMT responders than nonresponders, but these small subgroup comparisons were not definitive. Together, the directional findings provide a systems level context consistent with the plasma proteomic results.

Several features of this study limit how strongly the findings can be interpreted. The biomarker cohort was small, especially the sham and MMT nonresponder groups, and the response phenotype was defined post hoc for this study. Repeated neuropsychological testing may have introduced practice effects, although the sham-controlled design helps account for retest-related changes. The ELISA results support the SomaScan HHIP result, but the broader proteomic findings should be assessed in larger cohorts and with additional targeted assays. ORA identifies pathway level associations rather than direct pathway activation, and plasma measurements cannot determine which tissues produced the observed protein changes. The OrganAge findings provide directional systems level context but do not establish organ rejuvenation. The HHIP/SHH simulation is a conceptual model and does not estimate mechanism, dose, duration, or efficacy.

In summary, clinical response among MMT participants was associated with selective early plasma protein changes, lower inflammation and injury biology, higher repair and adaptive remodeling biology, and lower HHIP. HHIP was the leading protein associated with response among MMT participants, and the ELISA supported the SomaScan result. Together, these findings identify candidate response-associated biological changes that warrant additional validation and mechanistic investigation in larger studies.

## METHODS

### Study design and samples

This exploratory plasma proteomic analysis was conducted within a randomized, sham-controlled feasibility trial of MMT in participants with cognitive impairment associated with Long COVID. Paired baseline and week 4 plasma samples were analyzed from 30 participants, including 20 assigned to MMT and 10 assigned to sham. Clinical outcomes were collected at baseline, week 4, and week 8. The protocol was approved by the Program for the Protection of Human Subjects at the Icahn School of Medicine at Mount Sinai (STUDY-24-01276), and all participants provided written informed consent. The trial was registered at ClinicalTrials.gov (NCT06739668). Additional eligibility, device, adherence, and safety details are reported in the parent clinical study ^13^.

### Cognitive measures

As described ^13^, cognition was assessed across domains using a comprehensive neuropsychological battery, including the Wechsler Adult Intelligence Scale Fourth Edition (WAIS-IV) Digit Span (attention/working memory), Delis-Kaplan Executive Function System (D-KEFS) Color-Word Interference Test (processing speed/executive functioning), Trail Making Test (TMT) Parts A (processing speed) and B (executive functioning), Ruff 2&7 (processing speed), Symbol Digit Modalities Test (SDMT; processing speed), D-KEFS Verbal Fluency Test (language/fluency), Hopkins Verbal Learning Test–Revised (HVLT-R; verbal learning and memory), Brief Visuospatial Memory Test–Revised (BVMT-R; nonverbal learning and memory), Rey Complex Figure Test Copy (RCFT; visuospatial abilities), and Multilingual Naming Test (MINT; language/confrontation naming).

### Clinical response phenotype

Clinical response was evaluated using a post hoc clinician-selected response phenotype comprising five improvement-oriented clinical/cognitive components (Table 1). The outcome set was selected following discussion of candidate measures with a study clinician to represent a cross-section of cognitive and symptom domains. This phenotype was used as an exploratory grouping variable for downstream molecular analyses.

Week 8 response was defined exploratorily as meeting the clinically important change (CIC) threshold in at least 2 of 5 components. For each component, CIC required improvement from baseline to week 8 of at least 0.5 SD, calculated from the randomized cohort’s baseline distribution. The threshold served as a standardized exploratory criterion rather than a validated measure-specific minimal clinically important difference. Missing component values were counted as not meeting the CIC threshold. Responder rates were compared using a two-sided Fisher exact test. Component-level CIC distributions were summarized descriptively and compared between groups using two-sided Mann–Whitney tests (Supplementary Figure 1).

### SomaScan profiling and preprocessing

Peripheral blood was collected into EDTA tubes at baseline and week 4, processed to plasma, aliquoted, stored at −80 °C, and shipped on dry ice for proteomic profiling. Plasma proteins were measured using the SomaScan 11K platform ^15^. Following SomaLogic normalization and quality-control procedures, normalized RFU values were log2-transformed as log2(RFU) and analyzed as paired week 4 minus baseline deltas. The post-QC analysis set comprised 10,509 human ColCheck-pass assays.

### Exploratory MMT-associated feature screening

MMT-associated features were identified using a staged paired-delta screening workflow. First, week 4 minus baseline deltas were compared between MMT and sham using two-sided Welch tests. Second, within-MMT paired deltas were tested against zero using two-sided one-sample tests. Third, assays passing both screens in the same direction were retained as concordant MMT-associated features. For each screen, features were prioritized using unadjusted two-sided p ≤ 0.20 and absolute delta effect ≥0.10 log2(RFU) units. This liberal threshold was used for exploratory feature prioritization. The resulting set contained 69 SomaScan features corresponding to 66 unique protein targets.

### Exploratory 17 protein response pattern

The 69 concordant MMT-associated features were first reduced to one assay per unique protein using the treatment-screen evidence. The resulting 66-protein set was then intersected with the MMT responder versus nonresponder paired-change screen. The responder screen used an unadjusted two-sided p value threshold of 0.20 and an absolute responder minus nonresponder change difference of at least 0.10 log2(RFU), yielding the exploratory 17 protein response pattern. For composite scoring, protein changes were standardized across participants and direction aligned to the MMT responder versus nonresponder difference: values for proteins lower in responders were multiplied by −1, and values for proteins higher in responders were unchanged. The composite was calculated as the mean direction aligned standardized change, with higher values indicating greater similarity to the exploratory 17 protein response pattern. Heatmaps display standardized changes without direction alignment, preserving each protein’s observed direction of change.

### HHIP analysis

Hedgehog interacting protein (HHIP) change was defined as week 4 minus baseline log2 RFU for SomaScan assay 10833-64. Within the MMT group, responder versus nonresponder differences were tested using Welch tests across the full SomaScan feature set, with Benjamini-Hochberg adjustment across that testing family. The corresponding sham HHIP comparison also used a Welch test. HHIP separation within MMT participants was additionally evaluated using an exact two-sided label permutation test based on AUC distance from 0.5, along with bootstrap resampling and leave one participant out analyses. Because HHIP was selected and evaluated in the same small cohort, it was interpreted as an exploratory response associated marker.

### Targeted HHIP ELISA supporting analysis

To assess the SomaScan HHIP finding using a second assay method, we measured HHIP in baseline and week 4 plasma samples from participants who received MMT using the Reddot Biotech Human HHIP ELISA Kit (RDR-HHIP-Hu) and the manufacturer’s ELISA protocol. Concentrations were calculated from the standard curve. The manufacturer reported an assay range of 0.312–20 ng/mL and a sensitivity below 0.107 ng/mL. Week 4 minus baseline changes were summarized descriptively by response status to assess whether the direction of HHIP change was consistent with the SomaScan finding.

### Conceptual HHIP/SHH inflammation-resolution model

An unfitted conceptual model was used to explore whether repeated transient HHIP reductions could produce more persistent repair and inflammatory suppression than a single treatment week (Supplementary Figure 3). The model included normalized states for circulating HHIP H(t), unresolved damage D(t), and active inflammation I(t), with effective SHH/Hedgehog signaling S(t) and repair R(t) = 1 − D(t) as readouts. Its structure was motivated by literature supporting HHIP as a Hedgehog antagonist and context-dependent Hedgehog roles in inflammation and tissue repair ^21–24,29–31^.

Each modeled session transiently reduced circulating HHIP, which then returned toward baseline under SHH-induced HHIP negative feedback. Lower circulating HHIP was assumed to increase effective Hedgehog signaling. In the model, signaling above baseline accelerated reductions in unresolved damage and inflammation. No damage source or damage-regrowth term was included.

The model compared no treatment, one week with three sessions every 72 hours, and four weeks with eight sessions every 72 hours. The modeled timing was illustrative and used the minimum separation specified in the trial protocol. Conditions used the same 2,000 illustrative parameter sets and initial pulse draws. Under the model assumptions, the four week simulation showed greater reductions in unresolved damage and inflammation than the one week simulation.

The model was not fitted to participant level data, and its parameter sets are not virtual participants. The assumed effect of MMT on HHIP, plasma-to-tissue mapping, rates, and timing are conditional; the model does not estimate participant responses, pathway engagement, or efficacy.

### Directional overrepresentation analysis and protein signatures

Directional overrepresentation analysis (ORA) tested whether protein sets associated with MMT and clinical response converged on biological programs relevant to Long COVID, Neuro PASC, inflammation resolution, vascular injury, and repair biology. Literature derived signatures were curated from published Long COVID/PASC, Neuro PASC, immune profiling, vascular, endothelial, complement and coagulation, mitochondrial, oxidative stress, neutrophil, interferon, TNF, NFκB, HIF, and targeted proteomic studies ^7,8,32–39^. These signatures were analyzed separately from GO, KEGG, Reactome, and MSigDB gene sets, which were used as external annotation and sensitivity resources.

Increased and decreased protein sets were analyzed separately against the 9,343-gene measured background using one-sided hypergeometric tests. Gene sets containing 5–1,000 measured genes were tested; a minimum overlap of two genes was required for literature-derived signatures and three genes for external database signatures. Benjamini-Hochberg correction was applied within each source type, database, comparison family, and direction. ORA was used to summarize biological patterns rather than establish mechanism. Participant level program scores were calculated as the mean direction-aligned change z score across proteins assigned to the lower inflammation and injury and higher repair and adaptive remodeling programs, then compared between MMT responders and nonresponders.

### OrganAge analysis

OrganAge analysis was performed by Vero using its analytics. Pretrained plasma OrganAge models ^16^ were applied to week 0 and week 4 SomaScan profiles to estimate organ-specific and organismal OrganAge values. OrganAge change was calculated as week 4 minus week 0; negative values indicate a lower predicted age gap at week 4 than at baseline. Treatment effects were tested using age and sex adjusted linear models, and p values were FDR corrected across the 12 OrganAge signatures. The Brain and Organismal responder panels used direct two-sided Mann Whitney comparisons between MMT responders and nonresponders. The analyses were interpreted in light of the small sample and responder subgroup sizes.

### Statistical analysis and software

All tests were two-sided unless otherwise specified. Continuous comparisons used paired or between-group delta contrasts, Mann-Whitney tests, Welch tests, bootstrap procedures, leave one participant out analyses, permutation tests, and ROC/AUC summaries as appropriate. Multiple testing was controlled using Benjamini-Hochberg FDR correction where formal testing families were evaluated. Nominal protein-level p values were used for staged discovery prioritization, whereas FDR-adjusted q values were reported for pathway, OrganAge, and sensitivity analyses.

Analyses were implemented in Python using NumPy, pandas, SciPy, statsmodels, scikit-learn, matplotlib, and Jupyter-compatible notebooks. SomaScan ADAT files were parsed directly in Python, and assay-provided SeqId and target annotations were retained for feature mapping.

### AI-assisted code implementation

Analysis code and figures were developed in Python notebooks with assistance from OpenAI Codex. All results were verified and approved by the authors.

## Data Availability

All data produced in the present study are available upon reasonable request to the authors

## Acknowledgements

We thank the study participants for their time and commitment. We also thank the clinical research, neuropsychological assessment, phlebotomy, sample-processing, and data-management staff who supported participant visits, biospecimen collection, and study operations. We thank Devanshi Patel for logistical support with plasma proteomic profiling.

## Author contributions

N.R.B. and B.T.G. conceived the exploratory proteomic analysis. D.P., J.B. and B.T.G. designed the parent clinical study. D.M. contributed to the usability aspects of the study design. D.P. and J.B. oversaw participant recruitment, clinical assessments, and sample collection. N.R.B. performed SomaScan data processing, quality control, and statistical, clinical, pathway, and biomarker analyses. D.K. and W.Z. performed OrganAge analyses. N.R.B. and B.T.G. interpreted the clinical and molecular findings. N.R.B. drafted the manuscript. All authors critically revised the manuscript, approved the final version, and agree to be accountable for the work.

## Data availability

All data produced in the present study are available from the corresponding author upon reasonable request, subject to applicable institutional approvals, data-use agreements, and privacy protections.

## Code availability

No custom code or algorithms were developed for this study. Analyses were performed using publicly available Python packages as described in the Methods.

