## Supplemental Figures for "Plasma Proteomics Identifies a Microtesla Magnetic Therapy Response Signature in Long COVID"

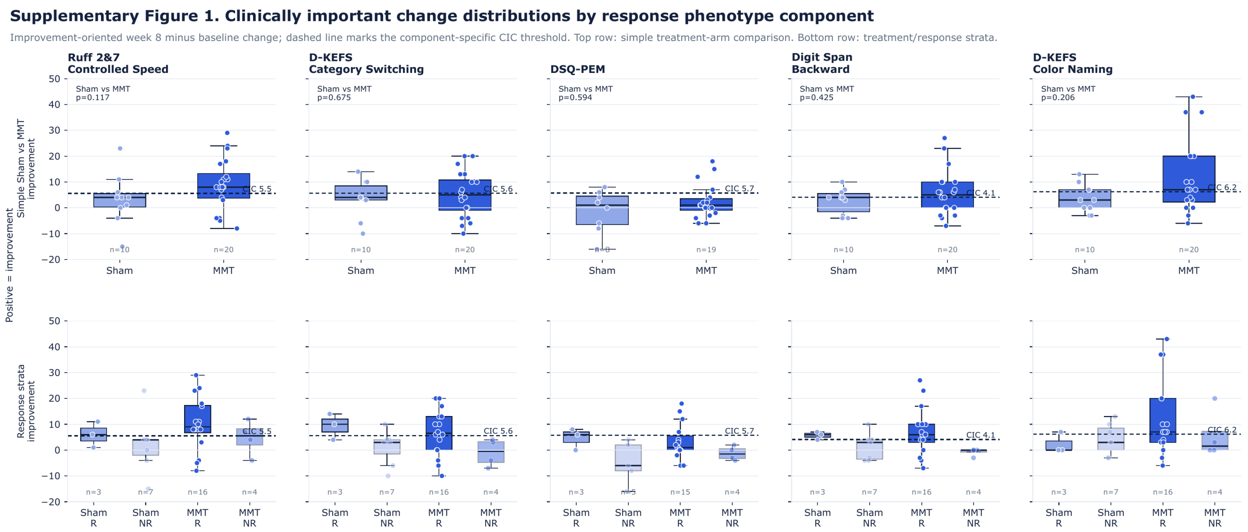


Supplementary Figure 1. Clinically important change distributions by response phenotype component. Component-level clinically important change (CIC) distributions are shown for the five response phenotype components: Ruff 2&7 Controlled Speed, D-KEFS Category Switching, DSQ-PEM, Digit Span Backward, and D-KEFS Color Naming. Week 8 minus baseline change was converted to an improvement-oriented score for each component, with positive values indicating clinical improvement. DSQ-PEM was direction-reversed because lower symptom burden indicates improvement. Dashed horizontal lines show component-specific CIC thresholds, defined as 0.5 baseline standard deviations of the corresponding measure. The top row shows simple sham versus MMT comparisons; the bottom row shows treatment/response strata. Two-sided Mann-Whitney tests were used for the simple sham versus MMT component comparisons.

**Supplementary Figure 2. Raw trajectories for the exploratory 17 protein response pattern**

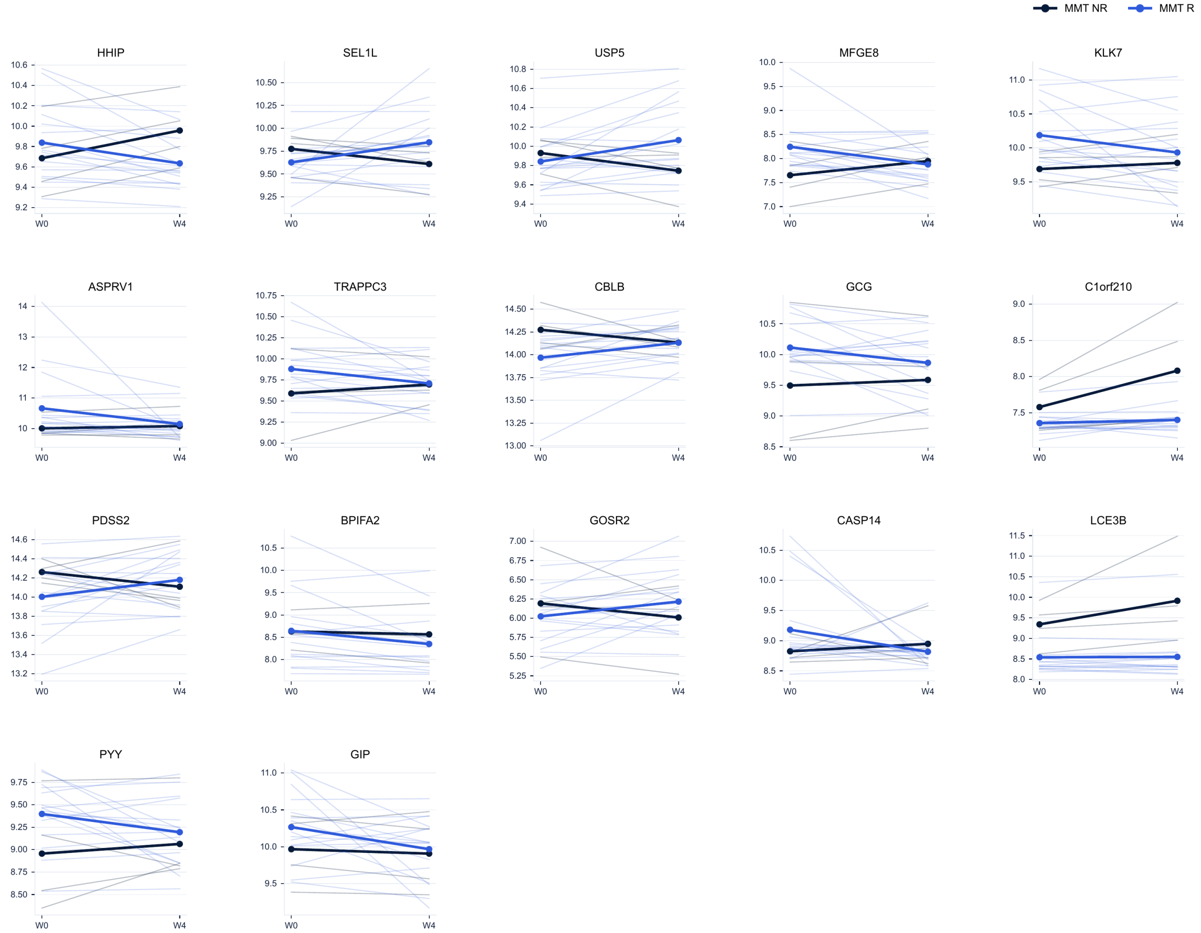


Supplementary Figure 2. Raw week 0 and week 4 trajectories for the exploratory 17 protein response pattern. Raw SomaScan trajectories are shown for MMT participants. Thin lines show paired participant values, and thicker lines show group mean trajectories for MMT responders and nonresponders. Values are displayed in each protein's observed direction before direction alignment for the composite. Several proteins showed different mean trajectories between responder groups, including HHIP, which decreased in MMT responders and increased in nonresponders.

### Supplementary Figure 3. Conceptual model for interpreting HHIP dynamics during MMT

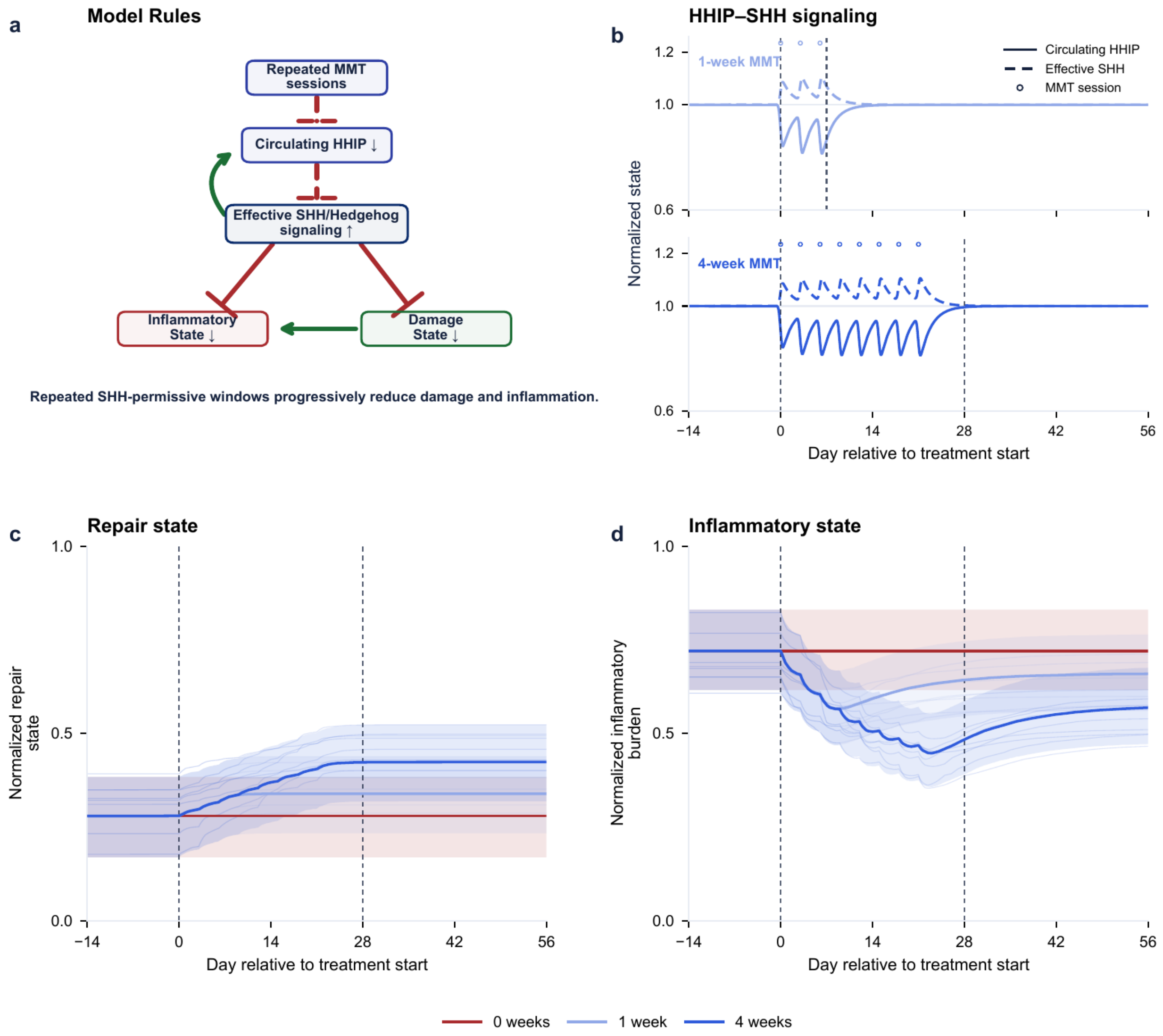


Supplementary Figure 3. Conceptual model for interpreting HHIP dynamics during MMT. a, Directed model rules. Dashed edges denote the hypothesized effect of MMT on HHIP and uncertain mapping from circulating HHIP to tissue SHH/Hedgehog permissiveness; solid edges denote internal model rules. b, Median circulating HHIP (solid) and effective SHH signaling (dashed); the inverse trajectories show the assumed signaling windows. c, Normalized repair, R = 1 − D. d, Normalized inflammatory burden. Curves show medians and shaded bands show 10th–90th percentiles across 2,000 paired illustrative parameter sets for no treatment, one week with three sessions every 72 hours on days 0, 3, and 6, and a four week condition with eight sessions every 72 hours on days 0, 3, 6, 9, 12, 15, 18, and 21. This matches the trial treatment schedule. Under the model assumptions, the simulation spanning four weeks showed greater damage removal and inflammatory reduction than the simulation spanning one week. The model is unfitted and does not estimate efficacy, represent participant responses, or establish that circulating HHIP measures CNS SHH/SMO/GLI activity.

### Supplementary Figure 4. HHIP ELISA supporting evidence

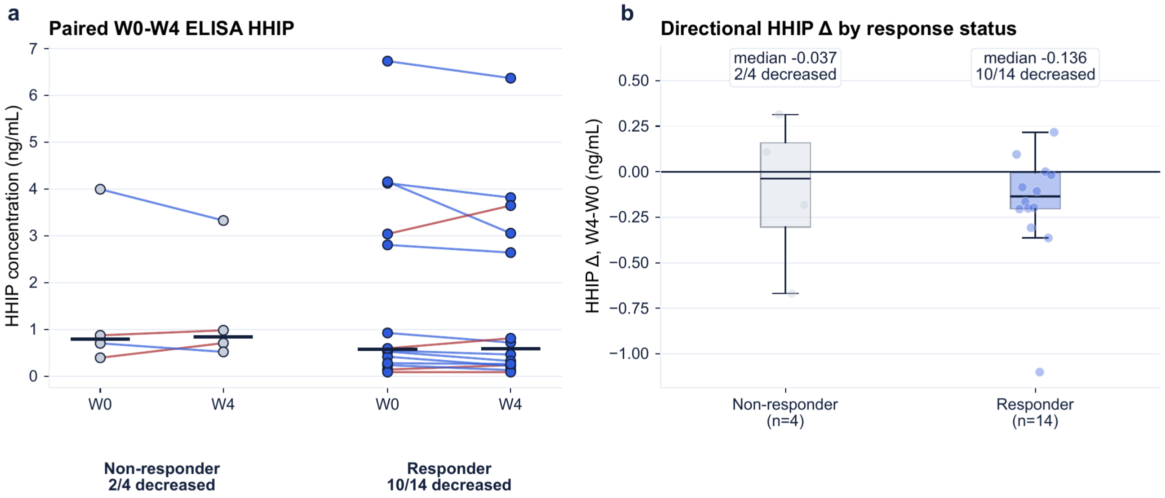


Supplementary Figure 4. Representative HHIP ELISA supporting evidence. a, Paired week 0 and week 4 plasma HHIP concentrations by response group. Lines show participant-level change; short horizontal bars mark group medians. b, Week 4 minus week 0 HHIP change by response group. In the displayed representative set, HHIP decreased in 10 of 14 responders and 2 of 4 nonresponders. The ELISA result was directionally consistent with the SomaScan HHIP pattern.

**Supplementary Figure 5. Directional ORA**

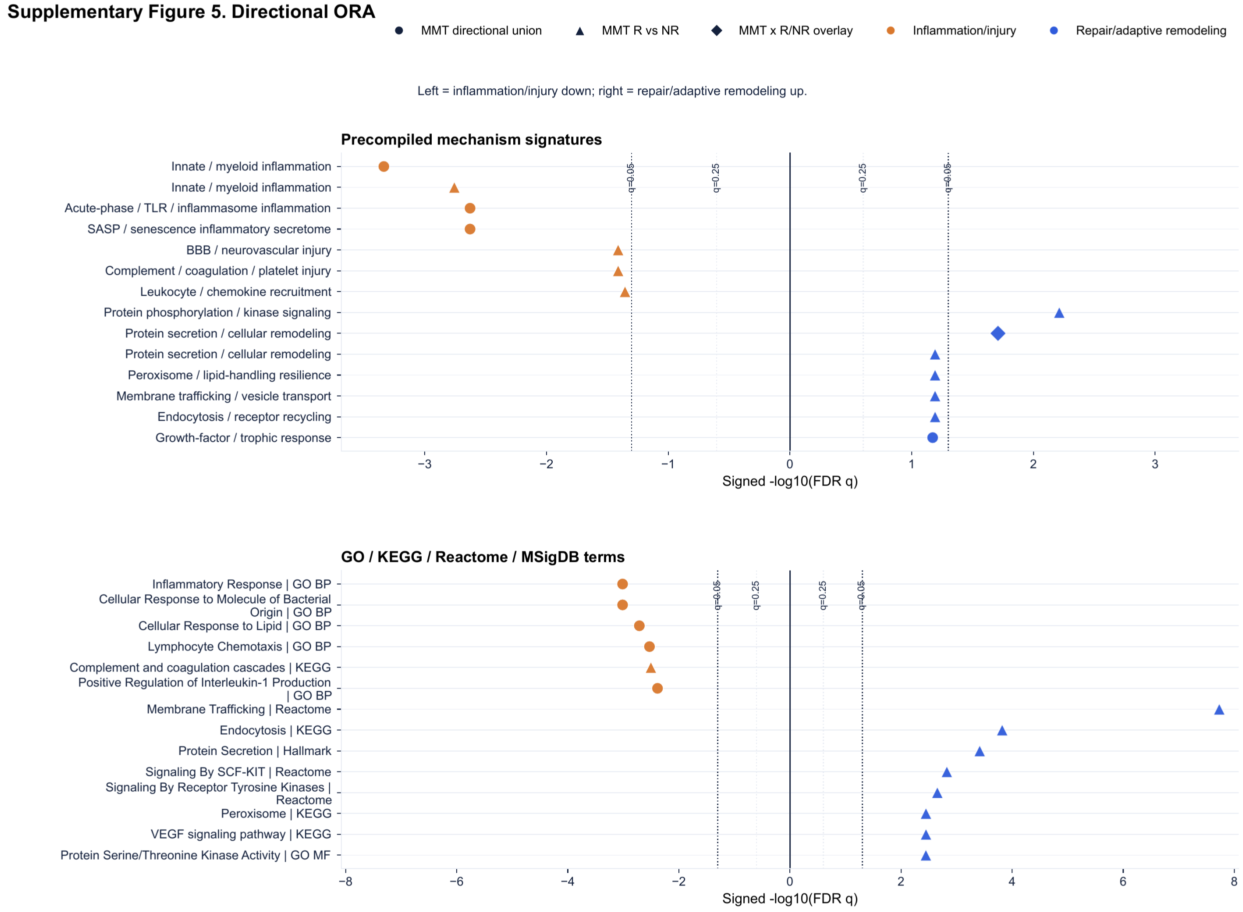


Supplementary Figure 5. Directional ORA. Directional ORA results are shown for mechanism-based signatures and public pathway and database gene sets. Each point represents a direction-consistent enriched biological term. Its horizontal position shows signed −log10(FDR q): negative values denote lower inflammation and injury biology, and positive values denote higher repair and adaptive remodeling biology. Greater distance from zero indicates stronger FDR-adjusted evidence. Point shape identifies the protein set used for enrichment testing. Dotted reference lines indicate q values of 0.05 and 0.25. The displayed terms provide detail on the biological programs summarized in Figure 6.

#### Supplementary Figure 6. Ranked enrichment sensitivity

Threshold-free enrichment using all measured SomaScan proteins; plotted terms are direction-consistent with the Figure 6 model.

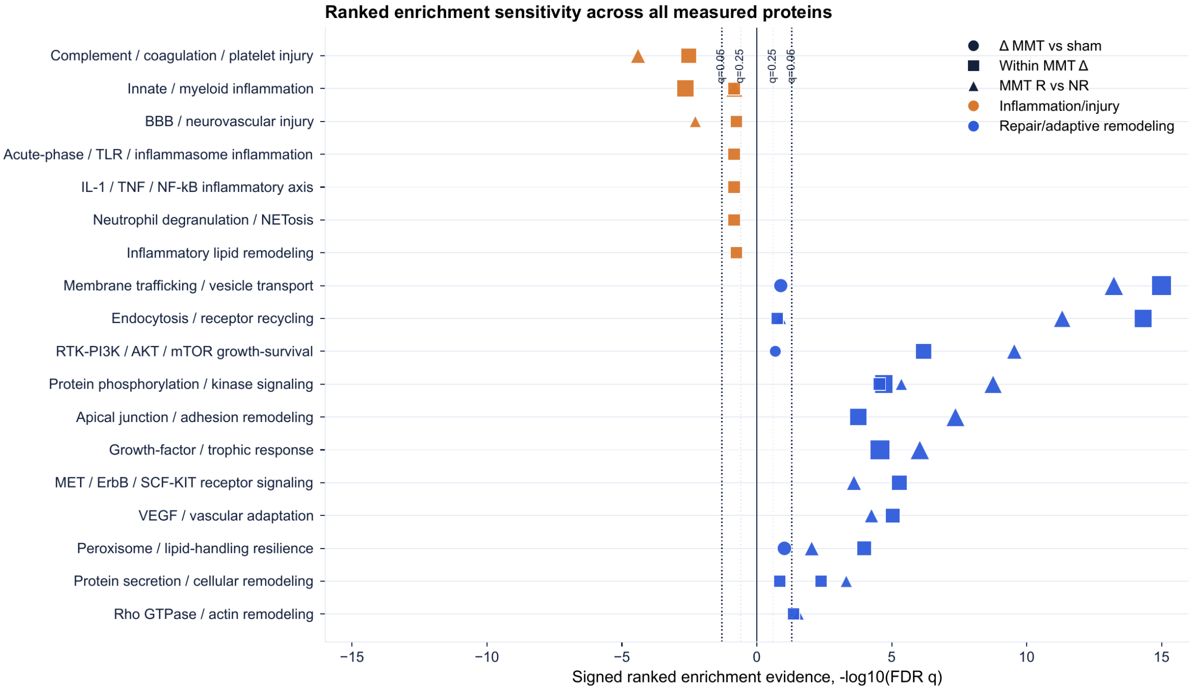


Supplementary Figure 6. Ranked enrichment sensitivity. Ranked enrichment provided a sensitivity analysis of the direction-aware ORA findings using all measured SomaScan proteins. Results are summarized by direction, source set, and false discovery rate-adjusted evidence. The ranked analysis was consistent with lower inflammation and injury biology and higher repair and adaptive remodeling biology in MMT responders compared with MMT nonresponders.
